# Natural language processing and modeling of clinical disease trajectories across brain disorders

**DOI:** 10.1101/2022.09.22.22280158

**Authors:** Nienke Mekkes, Minke Groot, Sophie Wehrens, Eric Hoekstra, Megan K Herbert, Maaike Brummer, Dennis Wever, Netherlands Neurogenetics Database Consortium, Bart J.L. Eggen, Annemieke Rozemuller, Inge Huitinga, Inge R. Holtman

## Abstract

Brain disorders, including neurodegenerative diseases and mental illnesses, are often difficult to diagnose and study due to clinical and pathological heterogeneity, overlap in clinical manifestations between disorders, and frequent comorbidities, hampering drug development and fundamental research. Hence, there is a clear need for data-driven approaches to disentangle these complex disorders. Here, we established a computational pipeline to process clinical summaries from donors with a wide range of brain disorders that were neuropathologically diagnosed by the Netherlands Brain Bank. First, we identified and defined 90 cross-disorder signs and symptoms within cognitive, motor, sensory, psychiatric, and general domains. Second, we trained and optimized natural language processing (NLP) models to identify these signs and symptoms in individual sentences of the extensive clinical summaries from donors of the NBB, resulting in temporal disease trajectories. Third, we studied the temporal manifestation and survival profiles across rare and complex dementias, alpha-synucleinopathies, frontotemporal dementia subtypes, and mental illnesses, giving new insight into how symptomatology differs in manifestation and temporal profiles across brain disorders. Lastly, we trained a recurrent neural network to predict the Neuropathological Diagnosis. Taken together, this integrated approach resulted in a highly unique resource that can facilitate research into cross-disorder symptomatology.

## Introduction

The brain is a highly complex organ that is susceptible to a wide range of disorders including neurodegenerative diseases, such as Alzheimer’s Disease (AD), frontotemporal dementia (FTD), Parkinson’s Disease (PD), neuroinflammatory disorders, such as Multiple Sclerosis (MS), and mental illnesses, including schizophrenia (SCZ), bipolar disorder (BP), and Major Depressive Disorder (MDD). Most brain disorders are inherently difficult to diagnose and study due to considerable heterogeneity^1–3^, shared clinical and pathological features between disorders^4,5^ and frequent and complex comorbidity patterns^6–8^. Moreover, large discrepancies between clinical and neuropathological diagnosis were frequently observed, in which up to a third of cases with a specific dementia were misdiagnosed^9–12^. The discrepancies between neuropathological diagnosis and clinical manifestations might be bridged by extensive clinical phenotyping and syndrome classification, in which the temporal manifestation of signs and symptoms is of crucial importance^3,7,13^. Temporal profiling also allows for overall survival analysis, that describes the rate of survival after a diagnosis was given or after a symptom was observed, which showed considerable differences between dementias. For example, after a clinical diagnosis of Dementia, patients with Dementia with Lewy Bodies (DLB) have considerably shorter lifespan than those with AD^14^. The survival profile of patients with rare and complex forms of dementia is currently largely unknown. Hence, there is a clear need for in-depth temporal profiling of signs and symptoms across brain disorders. Here, we aimed to delineate temporal signs and symptoms across brain disorders to improve differential diagnosis, and prognosis, and to address fundamental questions concerning the underlying neuropathological processes by mining the extensive clinical history data from donors of the Netherlands Brain Bank (NBB).

The NBB has performed almost 5000 human brain autopsies from donors with a wide range of brain disorders^15^. The NBB brain tissue is known for its ultra-short post-mortem delay, and extensive clinical and neuropathological characterization in the form of clinical/pathological summaries, making it a unique resource that has facilitated neuroscientific research globally. Specialized neuropathologists at the NBB perform extensive neuropathological examinations which, together with the clinical diagnosis result in a Neuropathological Diagnosis. These Neuropathological Diagnoses frequently deviate from the clinical diagnoses^9,10,12,16^. In order to study the symptomatology of these diagnosis we established an extensive computational pipeline consisting of parsers and Natural Language Processing (NLP) models to convert extensive clinical summaries from the NBB into standardized clinical disease trajectories, which allowed for temporal profiling, survival analysis, and predictive modeling of different brain disorders including alpha-synucleinopathies, rare and complex dementias, frontotemporal dementia subtypes and mental illnesses.

## Method and materials

### 2.1 NBB files

#### 2.1.1. NBB summaries

The NBB is a non-profit organization that supports human brain-research by providing well-characterized brain tissue from healthy and diseased individuals to the international scientific community, facilitating more than 100 scientific publications a year^17^. All adult citizens of the Netherlands can register to become donors in accordance with NBB procedures which are in full compliance with Dutch and European law. All NBB donors provided informed consent for their tissue and their data to be used for research purposes. The forms and procedures of the NBB were approved by the Free University Medical Center - Medical Ethics Committee (VUmc METC, Amsterdam, the Netherlands). Upon death of a donor, the NBB requested in-depth information from the medical specialists and general practitioner (GP)/geriatrician regarding the donor’s specific diagnoses, general health status, surgeries, and familial conditions. This information was summarized and translated from Dutch to English by trained medical staff under the auspices of the Coordinator Medical Information. These summaries are semi-standardized text files with free text fields to allow flexibility to add information from the medical specialist as seen fit. The files contain several headers such as ‘general information’, ‘clinical history’, ‘clinical diagnosis’, and ‘medication’. In the ‘clinical history’, signs and symptoms were described per year as they appeared in the medical information obtained, with minimal interpretation.

#### 2.1.2. NBB Neuropathological examinations and neuropathological diagnosis

After each brain autopsy, neuropathologists perform extensive macroscopic and microscopic neuropathological examinations for the NBB, of which the results are added to the clinical-neuropathological summaries as individual headers. Macroscopic information concerns the overall brain structure, such as brain atrophy, ventricle size, structure of the Circle of Willis, while the microscopic information is an in-depth characterization of many brain structures including the identification of protein aggregates and signs of cellular stress. The neuropathologists use this information to assign a final diagnostic label which we refer to as “Neuropathological Diagnosis” (Supplemental Table 2). This Neuropathological Diagnosis can contain either 1) a clearly defined neuropathological diagnosis such as AD, or 2) specific neuropathological traits that are not associated with a single medical diagnosis such as argyrophilic grains, or 3) a psychiatric diagnosis based upon clinical observations such as SCZ, 4) specific neuropathological traits combined that are final diagnostic labels used exclusively by the NBB, such as Dementia with Senile Involutive Cortical Changes (DEM-SICC) and Non Alzheimer’s dementia (NAD). Of note, DEM-SICC is dementia with neuropathological AD changes, but not sufficient to qualify as AD, while Non-Alzheimer’s dementia (NAD) is a form of dementia that does not fit into any of the other pathological criteria defined. 5) a neutral label such as ‘Control’, indicating the absence of or minimal changes in neuropathology. These ‘Control’ donors, however, did often suffer from other peripheral diseases, such as cancer.

Each donor can have multiple Neuropathological Diagnoses. For example, a given donor might have received both an AD and PD label if the neuropathological examination indicated both pathologies.

### 2.2. Parsing

The semi-structured clinical-neuropathological summaries were parsed using an extensive set of Python based parsers that identified specific headers, and synonyms of those headers, and captured all relevant information. Files that were not properly formatted were manually reformatted by adding the appropriate headers. We used fuzz token_sort_ratio from the FuzzyWuzzy library to identify these headers^18^, in which a positive match occurred when more than 95% of characters were matched to a reference list of all headers. Here, words in the sentence are alphabetically ordered, converted to lowercase, and punctuation is removed. The key headers that we aimed to identify were ‘clinical history’, ‘clinical diagnosis’, ‘microscopic neuropathology’, ‘macroscopic neuropathology’, ‘neuropathology conclusions’, and ‘general information’. Next, the ‘clinical history’ information was parsed per year, and per sentence, setting the stage for temporal profiling. Sentences without clear year descriptions were parsed into a ‘year unknown’ category. Other time labels, such as last 2 months, last two years, at birth, were converted into relevant years. Sentences with a ‘year unknown’ label and associated predictions were included in general data exploration but excluded from temporal profiling. Temporal descriptions that encompass a range of multiple years (e.g., 2005-2007) were manually converted to individual years (e.g., 2005, 2006, 2007). Sentences that cross-referenced to previous years, were manually adjusted (e.g., in comparison to 2003).

### 2.3 Selection of files from the NBB

Donors were selected based on sufficient clinical and neuropathological information, defined as the presence of more than 500 characters in the clinical-neuropathological summaries. The final selection consisted of 3,101 donors, with 199,901 sentences of clinical history data. These donors suffered from a wide range of brain diseases, and received one or multiple Neuropathological Diagnosis, from a list of 94 Diagnoses (Suppl. table 1, Suppl table 2). The most common Neuropathological Diagnoses and their numbers, age at death and sex-distribution were depicted in (Suppl. figure 1).

### 2.4 Defining signs and symptoms

To produce the final list of key clinical signs and symptom attributes (see Suppl. Table 3), we went through several iterations of identifying attributes and labeling individual sentences from the clinical history from a predefined random set of donors (Fig 1A, 1B). The list of signs and symptoms appearing in the clinical history was composed based on three criteria: 1. medical-scientific relevance, 2. sufficient presence in the ‘clinical history’, 3. unambiguity with respect to the definition. Clinical signs and symptoms used for the clinical diagnosis from the most common neurodegenerative and psychiatric disorders in the NBB were compiled. Additionally, signs and symptoms that are reflective of general well-being, health, and functioning were added. Ultimately, 90 clinical attributes were identified, optimized, and defined (including inclusion and exclusion criteria and examples) and externally validated by a licensed neurologist (Suppl. notes) that encompass 19 groupings, including ‘Disturbances in mood and behavior’, ‘Extrapyramidal signs’, ‘Cognitive and memory impairment’ in 5 broad domains: psychiatric, cognitive, motor, sensory/autonomic, general (Fig 1B).

**Figure 1.**
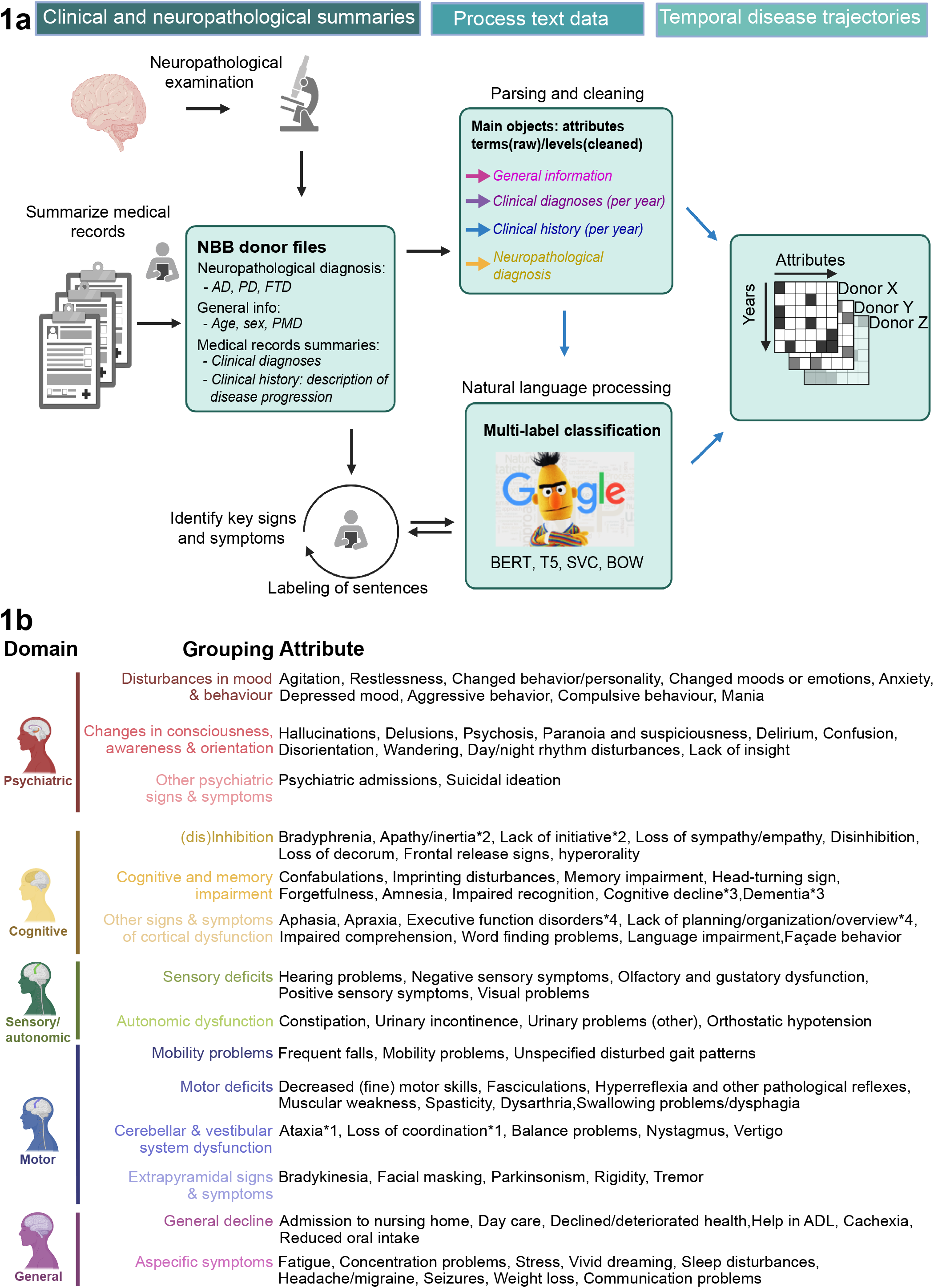
Introduction to the project. A) Workflow of the project. B) Signs and symptoms, their domains, and groupings, including colors and illustrative brain icons.

### 2.5 Labeling of donor files

Individual sentences from the ‘clinical history’ from a random predefined selection of donors were labeled according to whether the signs and symptoms were described. In total 293 donor files were selected, corresponding to approximately 10% of the data, to create a labeled training set for the final list of signs and symptoms. For each sentence, the signs and symptoms that were positively described were scored, resulting in a labeled dataset, containing 18,917 sentences. As mentioned above, multiple iterations of labeling were carried out on the ‘clinical history’ of the predefined randomly selected donor files, as the set of predefined signs and symptoms grew. In each iteration, (dis)concordant observations between scorers, and or the NLP models were discussed, further defined, examples and inclusion and exclusion criteria were given to signs and symptoms, and a new and larger set of sentences was scored. The final set of 293 donor files, containing 18,917 sentences, was labeled for the 90 attributes by one scorer.

### 2.6 NLP model optimization and comparison

The 18,917 labeled sentences were stratified and split for cross-fold validation (Figure 2A), for training and optimization of different NLP models. The Python library MultilabelStratifiedKFold (Bradberry (2018) was used to split the data into a test (20%) and training and validation (80%) fraction. The data was stratified to evenly distribute the different attribute labels over the test and training and validation sets^19^. The training and validation sets were split further using the same MultilabelStratifiedKFold library for the k-fold cross validation procedure used during model optimisation, with a k of 5. To ensure accurate comparisons, the same splits were used for the training and validation of every model.

**Figure 2.**
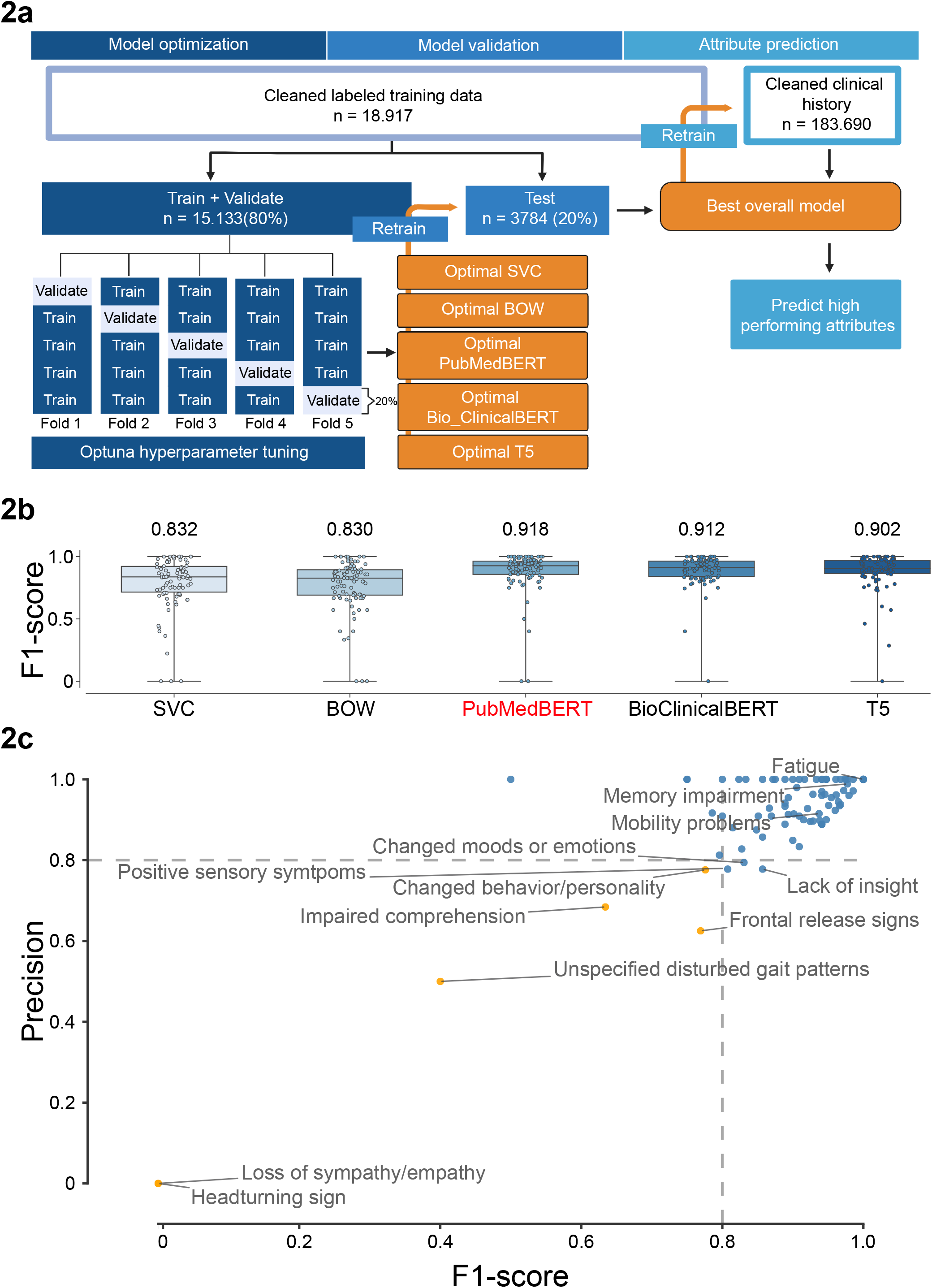
Natural Language Processing. A) Workflow of the NLP. B) Boxplots showing the F1-score per attribute, together with the average micro F1-precision. The best performing model, PubmedBert, was highlighted in red. C) Scatterplot depicting the classification performance of PubMedBERT in the form of precision and F1-score for individual signs and symptoms

Multiple multilabel classification NLP models were optimized and compared. We compared the performance of multiple NLP classification models on the labeled test data, to select the best performing model. The best performing model will be used to predict the entire clinical history. We selected two transformer models recently developed by Google called BERT^20^ and T5^21^. Different iterations of these models exist, mainly based on the contents of the pretraining corpora. We selected two pretrained BERT models: PubMedBERT^22^, Bio_ClinicalBERT^23^ that are optimized for medical text data. The standard version of the T5 model was selected. In addition, two common baseline models were used, a Bag Of Words model and a Support Vector Classifier in the form of linearSVC (Scikitlearn, Pedregosa *et al*., 2011). The Bag of Words model was implemented within a logistic regression framework on word frequency. Since both these models are not innately designed to be used for a multilabel problem, they were wrapped in the Scikitlearn package OneVsRestClassifier^24^. Optuna^25^ was used for hyperparameter tuning of all models by maximizing the average F1 score of the four cross-validation folds for 25 trials.

As our dataset is highly imbalanced, micro precision, recall and F1 score were used to evaluate model performance. Models were optimized with Optuna towards a high micro F1 score. As we value correct classifications (precision) more than correctly identifying every sentence (recall), we first identified the top 5 models based upon micro F1 score, from which we selected the final model based upon the highest micro precision score.

### 2.7 Descriptive statistics of symptom distributions among main-diagnosis groupings

#### 2.7.1 General statistics

The number of attributes that were labeled in the training set, and the number of predictions from the final model on the total set of all sentences were determined to calculate the attribute distribution. The year distributions were calculated from our parsed clinical files, to determine the period that the sentences extend, as well as the number of year observations and the number of sentences per year.

#### 2.7.2 Signs and symptoms distribution per main diagnosis

To identify signs and symptoms that were more frequently identified in specific disorders than expected, we compiled donors with the same diagnosis and set up a permutation test to test significant deviation from expectations. The total number of signs and symptoms were compiled for all donors with the same Neuropathological Diagnosis, and three statistics were calculated. First, the mean number of observations in sentences for donors belonging to a Neuropathological Diagnosis, and second the proportion of donors with a Neuropathological Diagnosis that contained minimally one observation of the symptoms. These results were plotted as a dot plot, in which the size corresponds to the proportion of donors for whom a sign or symptom was observed, and the color corresponds to the mean number of observations. The color-cut-off was set to a max of 5. The figure also contains a highlighted gray circle around the dot that indicates whether the sign or symptom was expected in the specific disorder, given a specific diagnosis. An asterisk was depicted if the attribute was more commonly observed than expected given a random background distribution as calculated with a permutation test. The random background distribution was calculated by randomly permuting the diagnosis labels of the individual donor data with 100,000 permutations. The p-value was calculated as the proportion of observations in which the observed value was higher than the random background, and was multiple testing corrected using the Benjamini Hochberg - False Discovery Rate. Moreover, we performed a chi-square test to identify whether the significant attributes (asterisk) per main diagnosis and the clinically expected attributes (circles) were overrepresented. The background consisted of an equal number of donors, who were randomly selected from all the donors belonging to main-diagnosis-categories, from which donors with the diagnosis of interest were removed.

Donors were compiled and studied according to subsets of neuropathological disorders. First, we compiled donors with the most common single Neuropathological Diagnosis. Second, we compiled frontotemporal dementia (FTD) subtypes, including Pick’s Disease, FTD-TDP-A (associated with progranulin mutations), FTD-TDP-B (associated with C9ORF72 mutations) FTD-TDP-C (neuropathologically characterized by predominantly long dystrophic neurites with rare neuronal cytoplasmic inclusions), and associated conditions, including Amyotrophic Lateral Sclerosis (ALS), Corticobasal Degeneration (CBD), Progressive supranuclear palsy (PSP). Third, we compiled rare and complex dementias (including those with complex comorbidities). Last, we compiled donors with psychiatric labels including, SCZ, MDD, BP, Obsessive Compulsive Disorder (OCD), Autism Spectrum Disorders (ASD).

#### 2.7.3 Observational profiles of the signs and symptoms

To test whether the number of year observations differed between different Neuropathological Diagnoses, we calculated the distribution of the number of year observations per donor within each Neuropathological Diagnosis and performed pairwise Mann-Whitey tests with Scipy, followed by a False Discovery Rate (FDR) multiple Testing Correction. These results were visualized as a Seaborn violin plot that is accompanied by a heatmap showing the results of pairwise significance testing, with -10log FDR corrected p-values depicted in orange when significant (p <= 0.01) (Figure number?).

#### 2.7.4 Temporal profiles of the signs and symptoms

To test whether the distribution of observations of a given sign or symptom differed temporally between disorders, we performed pairwise Mann-Whitey tests with Scipy, followed by a False Discovery Rate (FDR) multiple Testing Correction. These results were visualized as a Seaborn violin plot. Each violin plot is accompanied by a heatmap showing the results of pairwise significance testing, with -10log FDR corrected p-values depicted in orange when significant (p <= 0.01). These results were also plotted as a kernel density plots depicting the distribution of the temporal observations across all donors compiled according to their main diagnosis were made with Seaborn Displot, kne type, with age along the X-axis^26^. To normalize the numbers of donors with a diagnosis, all observed years in which an attribute occurred were added to a list. Random values were drawn from this list, proportional to a specified set of donors (N=100).

#### 2.7.5 Survival analysis

Survival analysis plots depicting the survival of the patients after the first observation of a given trait were made with Scikit Kaplan Meier Estimator. To test whether the survival after the observations of a given sign or symptom differed temporally between disorders, we performed pairwise Mann-Whitey tests with Scipy, followed by a FDR multiple Testing Correction. These results were visualized as a Seaborn violin plot. Each violin plot is accompanied by a heatmap showing the results of pairwise significance testing, with -10log FDR corrected p-values depicted in orange when significant (p <= 0.01).

#### 2.7.6 Cumulative proportion plots of psychiatric traits in psychiatric conditions

For donors with SCZ, BP, MDD, and controls, the number of year observations with specific relevant traits were calculated, converted into proportions for each diagnosis, and plotted as a stacked bar plot with the Pandas function plot, type = ‘stacked’. Symptoms that were observed more than 5 times were merged in the 5 observations categories. For each diagnosis, the proportion of donors with the number of year observations were calculated and plotted up to 5 year observations.

## Results

### 3.1 Identification of signs and symptoms and labeling of the data

To obtain temporal disease information on cross-disorder signs and symptoms, we have set up a computational pipeline that consists of text-parsers and an optimized NLP model to process extensive clinical summaries from donors with a wide-range of brain diseases from the NBB (Fig 1A, Suppl. Fig 1A). First, we identified a novel cross disorder clinical categorization system that contains 90 clinical signs and symptoms, including 4 synonyms signs and symptoms, associated with brain disorders and overall well-being/functioning that we subdivided into 14 groupings such as ‘Disturbances in mood & behavior’, ‘Cerebellar & vestibular system dysfunction’, ‘Memory impairment’, ‘Autonomic dysfunction’ and 5 broad domains: ‘Psychiatric’, ‘Motor’, ‘Cognitive’, ‘Sensory/autonomic’, and ‘General’ (Fig 1B). For each sign and symptom, we formulated the medical definition, and inclusion/exclusion criteria in relation to existing literature (Suppl. notes, table 1).

In order to identify whether these 90 signs and symptoms were positively stated in individual sentences, we labeled 18,917 sentences from a random set of 293 donors to train and validate different NLP models (Suppl. Fig 1B). Importantly, the data was highly imbalanced, in which most sentences had no attributes associated with them (Suppl. Fig 1C). Additionally, the number of observations for each label varied from 6 observations of ‘Fasciculations’ which were only observed in donors with Motor Neuron Disease (MND) to 466 observations of ‘Dementia’ which were mostly observed in patients with AD, Parkinson’s Disease with dementia (PDD), DLB, Vascular Dementia (VD), FTD (Suppl. Fig 1D). Next, we performed an enrichment analysis to determine whether a sign or symptom is more frequently observed in each disorder than expected by random chance. This analysis identified many expected disease specific signs and symptoms such as ‘Dementia’ being significantly enriched in AD, PDD, DLB, and VD but not in PD without dementia (Suppl. Fig 1D) and ‘Bradykinesia’ in PD, PDD, multiple system atrophy (MSA), PSP. These observed signs and symptoms were significantly overrepresented for *a priori* defined, and medically expected signs and symptoms (χ^2^= 171.28, p=1E-31).

### 3.2 NLP training, optimizing and predictions

To reliably identify these signs and symptoms in individual sentences, we established a pipeline to train, optimize and compare different NLP model architectures (Fig 2A). Firstly, the data was divided into a training and hold-out test set, stratified according to a relatively equal distribution of labels. We then employed a stratified 5-fold cross validation approach, where models were trained in 4 folds, and validated on the remaining part of the data. Five different model architectures (bag of words, Support vector classifier, Bio_ClinicalBERT, PubMedBERT, and T5) were trained and optimized with Optuna, and the best performing models, according to average micro F1-score (Suppl. figures 2A, 2D, 3A, 3D, 3G) and average micro precision (Suppl. figures 2B, 2E, 3B, 3E, 3H) were selected. Almost all signs and symptoms were reliably identified by all models, but a small subset performed considerably less well (Suppl. figures 2C, 2F, 3C, 3F, 3I), which consistently included the same signs and symptoms, including ‘Loss of Sympathy / Empathy’, ‘Head turning sign’, and ‘Unspecified disturbed gait pattern’. Secondly, the highest scoring iterations of each model architecture were compared using the hold-out test data, on which PubmedBERT showed the best model performance (Figure 2B). Thirdly, the optimal PubmedBERT architecture was retrained on all labeled data for the prediction of the 84 signs and symptoms that exhibited a micro precision >= 0.8 or a micro F1-score= > 0.8 (Fig 2C). Fourthly, this final model was used to predict whether specific signs or symptoms were described in individual sentences of the full corpus of individual sentences that were parsed per donor per year. For the final temporal profiling dataset, the four synonyms were merged, and the predictions of multiple sentences per year were collapsed on year level, to determine whether a sign or symptom was observed each year, as the minimal unit.

### 3.3 Interpretation of the unique datasets

The NLP models resulted in a unique dataset of signs and symptoms that were observed by the model in individual sentences that were parsed per year per donor. Again, we compiled donors according to their diagnosis, while only focusing on donors with a single Neuropathological Diagnosis and ran a permutation test to determine significance (Fig 3A). In this dataset, 269 signs and symptoms were significantly enriched in specific diagnoses, of which 148 were also *a priori* expected, a highly significant enrichment (χ^2^=295.96, p*=*2.5e-66). Importantly, the enrichment of the predicted dataset is much more significant for clinically expected signs and symptoms than the enrichment of the smaller labeled dataset, offering orthogonal evidence for the validity of our NLP approach.

**Figure 3.**
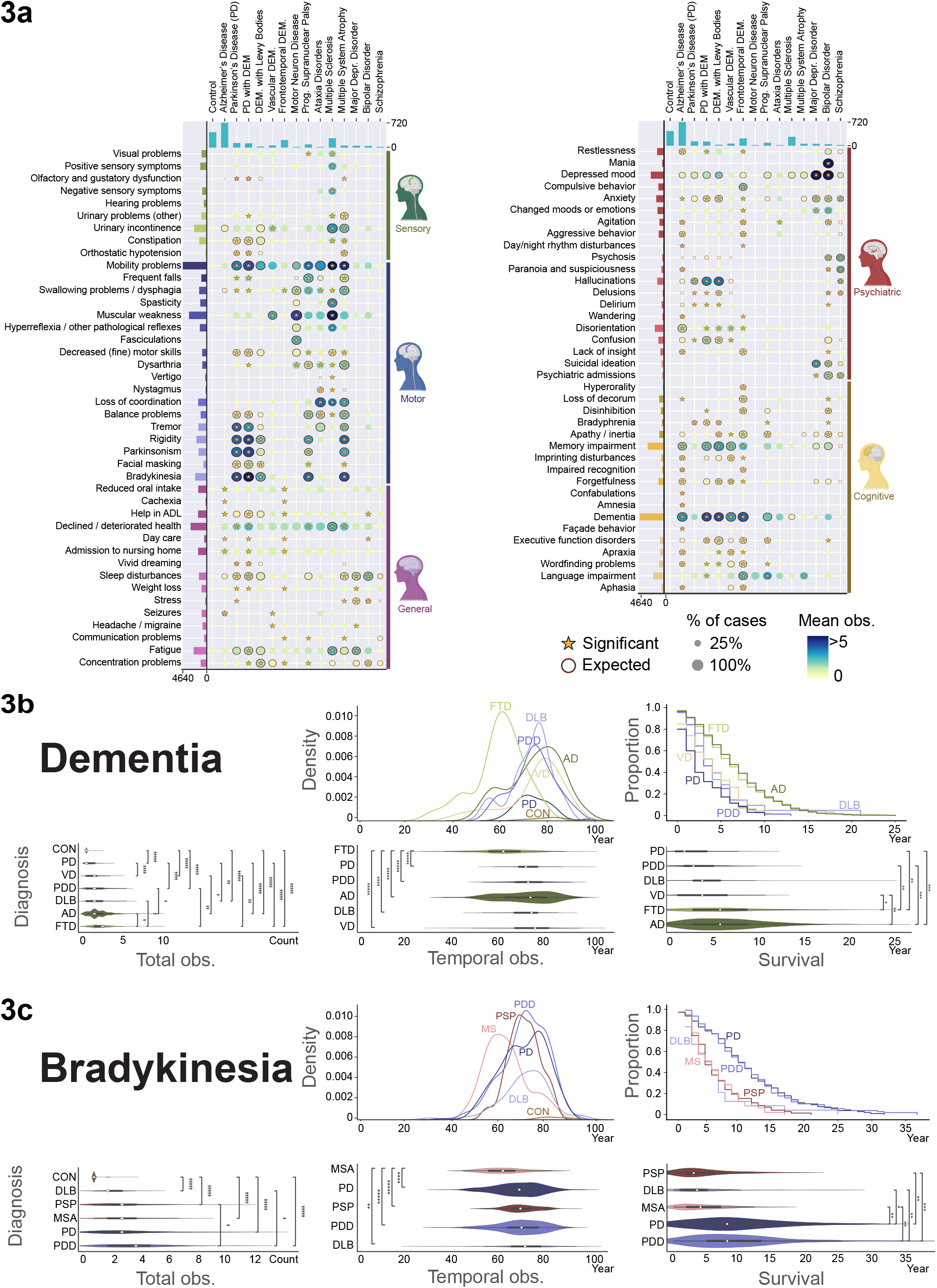
Interpretation of the model predictions. A) Dot plot and significance testing of signs and symptoms across disorders for the predicted dataset. The dot plot contains information about the proportion of donors in which an attribute was observed, corresponding to the size of the dot. The mean number of observations corresponding to the color of the dot. Significance, asterisk, is based upon a permutation test, with an FDR corrected P < 0.1. Orange surrounding circles highlight signs and symptoms that, *a priori*, were medically expected to be observed given a diagnosis. B) Temporal profiling plots of ‘Dementia’ for relevant disorders (AD, PD, PDD, DLB, VD) showing a seaborn density plot, a Kaplan Meier plot, and 3 Violin plots with significance testing (FDR-corrected P -values). *: p <= 1.00e-02, **: 1.00e-06 < p <= 1.00e-04, ***: 1.00e-08 < p <= 1.00e-06, ****: 1.00e-10 < p <= 1.00e-08, *****: p <= 1.00e-10. C) Temporal profiling plots of ‘Bradykinesia’ for all relevant disorders (PD, PDD, DLB, MSA, PSP). All figure plots are identical to B.

Interestingly, all included signs and symptoms were significantly enriched in at least one brain disorder, suggesting that all the signs and symptoms we identified were uniquely relevant for a subset of disorders. ‘Dementia’ was the most observed sign, followed by ‘Mobility problems’, ‘Muscular weakness’, ‘Memory impairment’, ‘Declined deteriorated health’, and ‘Depressed mood’, suggesting that these attributes were of particular importance for disease progression in this cohort. Of note, the sign and symptoms distribution in the predicted dataset strongly mirrored the labeled dataset. No signs and symptoms were significantly enriched in controls. As expected, ‘Dementia’ and ‘Memory problems’ were significantly enriched in dementias including AD, FTD, DLB, VD, PDD, but not in PD without dementia. Similarly, MS showed a striking enrichment for ‘Mobility problems’, and ‘Muscular weakness’, and ‘Fatigue’, which is very much in line with its disabling pathology of the brain and spinal cord. However, where ‘Mobility problems’ were significantly enriched in MS, PD, PDD, PSP, and MSA, ‘Muscular weakness’ was enriched in VD, MND, and MS, showing that our approach is able to detect a unique compendium of signs and symptoms in a disorder specific manner. Similarly, DLB had considerably fewer observations of extrapyramidal signs and symptoms such as ‘Bradykinesia’ ‘Facial masking’, ‘Tremor’ and ‘Rigidity’ than PD, and PDD, while many of these signs were also enriched in PSP and MSA, two conditions that are frequently misdiagnosed with PD (Koga, 2015; Owalabi, 2013). Moreover, major depressive disorder (MDD), and bipolar disorder (BP) showed a striking enrichment for ‘Depressed mood’, a key symptom in both disorders, while donors with BP also exhibited a strong enrichment for ‘Mania’. These findings suggest that we retrospectively have created a unique dataset that describes the clinical signs and symptoms that are associated with a large range of CNS disorders.

Next, we aimed to use this dataset to study the temporal profiles of specific signs and symptoms across disorders. To this end, we calculated three different statistics. First, we calculated the total number of year-observations in each condition in relation to the donors, to determine whether specific signs and symptoms were significantly more frequently observed in different diagnoses. Second, we calculated the temporal profile of those signs and symptoms, as a distribution of the years in which they were observed. Third, we performed a survival analysis to determine whether there are differences in the overall survival rate after the first observation of a sign or symptom between donors with different Neuropathological Diagnosis. As expected, the total number of observations of ‘Dementia’ was lowest in control donors, and highest in AD and FTD (Fig. 3B). Temporally, we found that ‘Dementia’ was observed significantly earlier in FTD than in other dementias, a well-known fact about FTD^27^. Interestingly, the survival analysis showed that after the first observation of ‘Dementia’, the survival of donors with PDD, PD, DLB and VD was significantly shorter than donors with AD, and FTD. PD, PDD, DLB are referred to as alpha-synucleinopathies, neurological conditions that are characterized by alpha-synuclein aggregation, resulting in neurodegeneration, frequently associated with dementia. Our findings suggest that ‘Dementia’ in the alpha-synucleinopathies is associated with faster disease progression than in AD, or FTD.

Next, we performed the same analyses with ‘Bradykinesia’, which showed that ‘Bradykinesia’ was significantly less frequently observed in DLB than in PD, PDD, and MSA. MSA is also an alpha-synucleinopathy, which primarily manifests by motor disturbances without cognitive and or psychiatric difficulties. There is considerable debate whether these alpha-synucleinopathies are different manifestations of the same underlying neuropathology manifesting in different brain regions, or that there are unique neuropathological processes associated with these disorders^6^. Temporally, we found that ‘Bradykinesia’ was observed significantly earlier in MSA than in the other disorders. Contrarily, the survival analysis showed that donors with MSA, PSP, and DLB with ‘Bradykinesia’ had significantly shorter survival than donors with PD, and PDD. These findings are in line with the hypothesis that there are qualitatively different aspects to these disorders, in which PD, and PDD share most features, but that DLB, and especially MSA are uniquely different.

Next, we extended the temporal analysis of ‘Dementia’ to other rare and complex forms of dementia including the combined diagnoses AD-PD, AD-DLB, AD-Vascular Encephalopathy (AD-VE), Dementia with Argyphilic grains (DEM-ARG), Dementia with Senile Involutive Cortical Changes (DEM-SICC), in which the pathology resembles AD, but does not qualify according to all neuropathological criteria, Dementia with Lewy Bodies and Senile Involutive Cortical Changes (DLB-SICC), Non Alzheimer’s Dementia (NAD), Alzheimer’s Disease with congophilic angiopathy (AD-CA). We ranked these dementias according to the median observation of ‘Dementia’ across time, and again found that ‘Dementia’ was observed the earliest in FTD. However, in many rare and complex dementias, including DEM-VE, DEM-SICC, AD-VE, ‘Dementia’ was observed at significantly later age than AD and VD (Suppl. Fig 4A), suggesting that the pathogenesis strikes at later age in these disorders. Interestingly, the observations of ‘Dementia’ in donors with both AD-PD were significantly later than donors with AD, PD, DLB and PDD. However, the survival analysis after the first observation of ‘Dementia’ showed a reverse profile, in which donors with AD-PD have significantly shorter overall survival after the first observation of ‘Dementia’, which was very similar to PD in general, while donors with AD, FTD, and AD-DLB have the longest survival. Interestingly, many of these temporal and survival profiles were also corroborated when the analysis was based on ‘Disorientation’ and ‘Memory impairments’, showing the robustness of the observed temporal and survival profiles.

Next, we explored the temporal profile and survival of several other prominent motor signs including ‘Mobility problems’ and ‘Muscular weakness’ (Suppl. Fig 4B). Where ‘Mobility problems’ were most uncommon in MND and frequently observed in PD, and PDD, ‘Muscular Weakness’ was most uncommon in PD, PDD, and much more common in MND. Both signs were observed at earliest time points in MS, followed by MND and MSA. Interestingly, PSP showed ‘Muscular weakness’ much later than ‘Mobility Problems’. These analyses corroborate the notion that many brain disorders exhibit partially overlapping clinical symptoms that manifest in a temporal fashion, potentially indicative of the neuronal substructures that are primarily affected.

Next, we partitioned donors according to FTD subtypes (Fig 4A). FTD is a group of highly heterogeneous conditions, in which degeneration of the frontotemporal cortex is a common hallmark^27, Staffaroni, 2022^. Different familial mutations and subtypes were identified including FTD-Fused in Sarcoma (FTD-FUS), FTD-TAR-DNA binding protein -43 (FTD-TDP), with its subtypes FTD-TDP-A (association with Progranulin mutations), FTD-B (association with C9ORF72 mutation), and FTD-C (characterized by predominantly long dystrophic neurites with rare neuronal cytoplasmic inclusions) (Mackenzie, 2011), and other related FTD conditions including Pick’s disease, PSP and Corticobasal degeneration (CBD). Again, ‘Dementia’ was the most observed sign in this FTD cohort, which, as expected, did not show in ALS (Fig 4A). ‘Dementia’ was observed significantly higher in all subtypes compared to control and was significantly lower in ‘PSP’ cases than in other FTD subtypes, suggesting that this FTD subtype is less affected by dementia (Fig 4B). Temporally, ‘Dementia’ was observed earliest in FTD-FUS, CBD, and FTD-TAU, and latest in PID, FTD-TDP-C and PSP. This temporal profile was consistent when these analyses were performed using ‘Memory impairment’. Next, we checked for signs and symptoms that were significantly different between specific FTD subtypes and found that ‘Compulsive behavior’ was consistently higher in TDP-B, TDP-C compared to many other FTD subtypes. Many of these observations were in line with and extended upon earlier work and can contribute towards a better understanding of the relationship between neuropathology and clinical syndromes in these disorders^28^.

**Figure 4.**
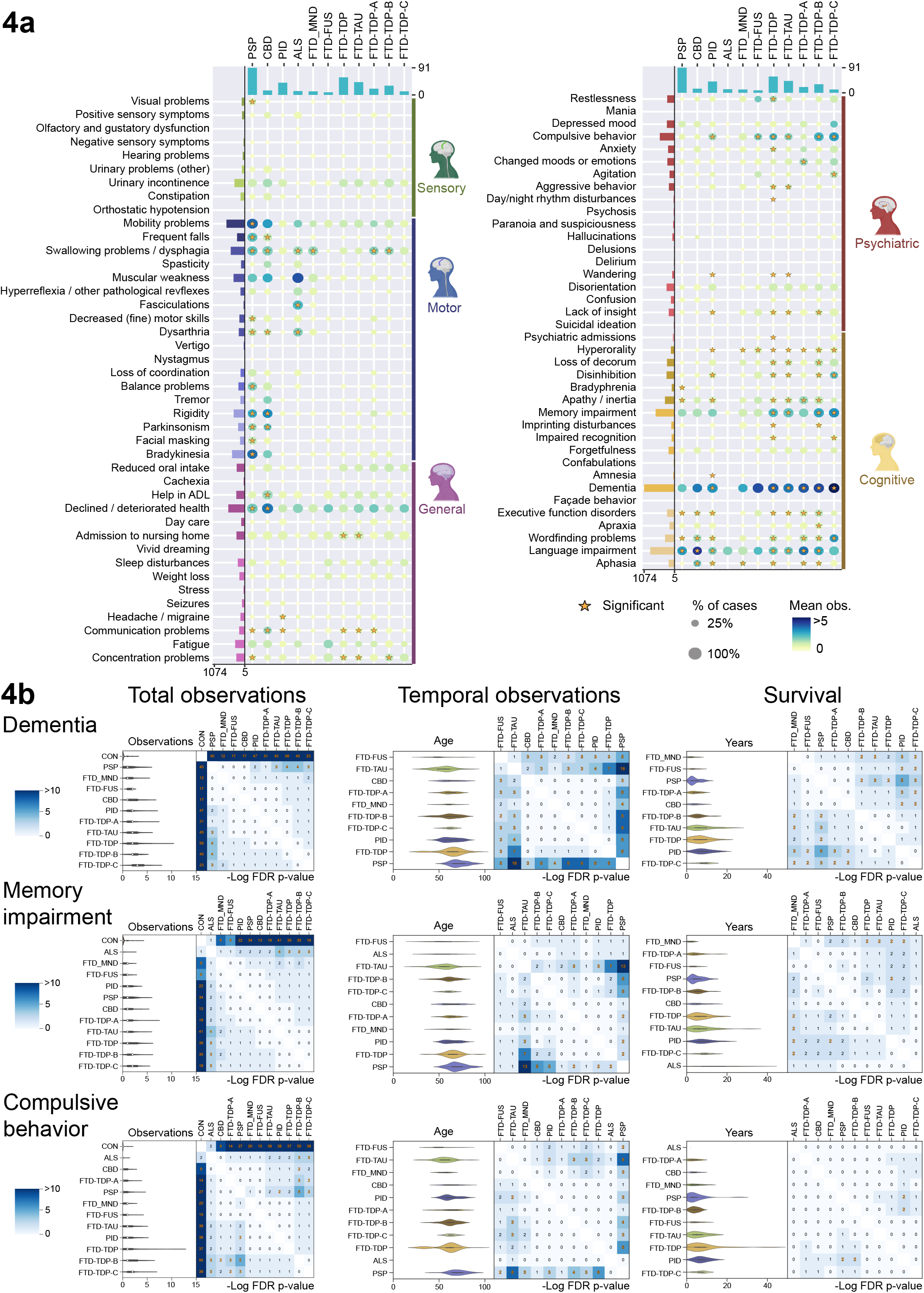
Analysis of the FTD subtypes. A) Dot plot and significance testing of signs and symptoms across FTD subtypes as described in 3A. B) Violin plots depicting the observation distributions, the temporal distribution, and survival distributions of ‘Dementia’, ‘Memory Impairment’, and ‘Compulsive Behavior’ Each violin plot is accompanied by a heatmap showing the results of pairwise significance testing, with -10log FDR corrected p-values depicted in orange when significant (p <= 0.01).

Lastly, we partitioned donors according to mental illnesses (Suppl. Fig 5A). As expected, donors with obsessive compulsive disorders, and autism showed an enrichment for ‘Compulsive behavior’, while ‘psychiatric admission’ and ‘suicidal ideation’ were particularly common in post-traumatic stress disorder. Increasing lines of evidence suggest that mental illnesses are not discrete categories but that individuals suffering from these disorders’ manifest behavior along a spectrum of traits^29^. To test this hypothesis, we looked at the most common symptoms in this dataset and plotted the number of observations as a cumulative proportion (Suppl. Fig 5B). ‘Mania’ was largely confined to donors with BP, especially for donors with more year-observations. Similarly, ‘Depressed mood’ was largely confined to donors with BP and MDD but to a lesser degree also to SCZ. For many other symptoms much more fluidity was found, for example ‘Hallucinations’ were more common in SCZ, but a relatively large subset of donors with SCZ didn’t manifest this trait at all. Moreover, ‘Psychosis’, ‘Anxiety’, and ‘Fatigue’ showed clear increases in all three disorders compared to controls. These observations are in line with the hypothesis that mental illnesses are not discrete disorders, but a continuum of signs and symptoms that manifest temporally.

### 3.4 Predicting brain disorders using signs and symptoms

We next aimed to build a predictive machine learning model using this unique temporal dataset to predict the underlying Neuropathological Diagnoses from temporal signs and symptoms. We implemented a gated recurrent unit (GRU-D) that is particularly well suited to work with time-series data with missing values, as often encountered in health care^30^. Again, we established a workflow to train, optimize and predict the Neuropathological Diagnosis from temporal signs and symptoms (Fig 5A). As expected, the model showed clear improvement over successive epochs, while no signs of over-fitting emerged (Fig 5B). This model was able to reliably diagnose most donors and showed errors that are comparable to medical doctors (Fig 5C, Fig 5D). For example, the most frequent misdiagnoses were observed between PD, and PDD; and VD, and AD; and SCZ, MDD and controls, which are also frequently clinically misdiagnosed. It is important to note that the numbers of donors in the disorders also correlated considerably with the diagnostic ability of our model, which suggest that a larger sample size, and or improved modeling approaches are necessary for obtaining clinical relevance. This study could function as a steppingstone towards improved clinical diagnosis of complex brain disorders at earlier time points.

**Figure 5.**
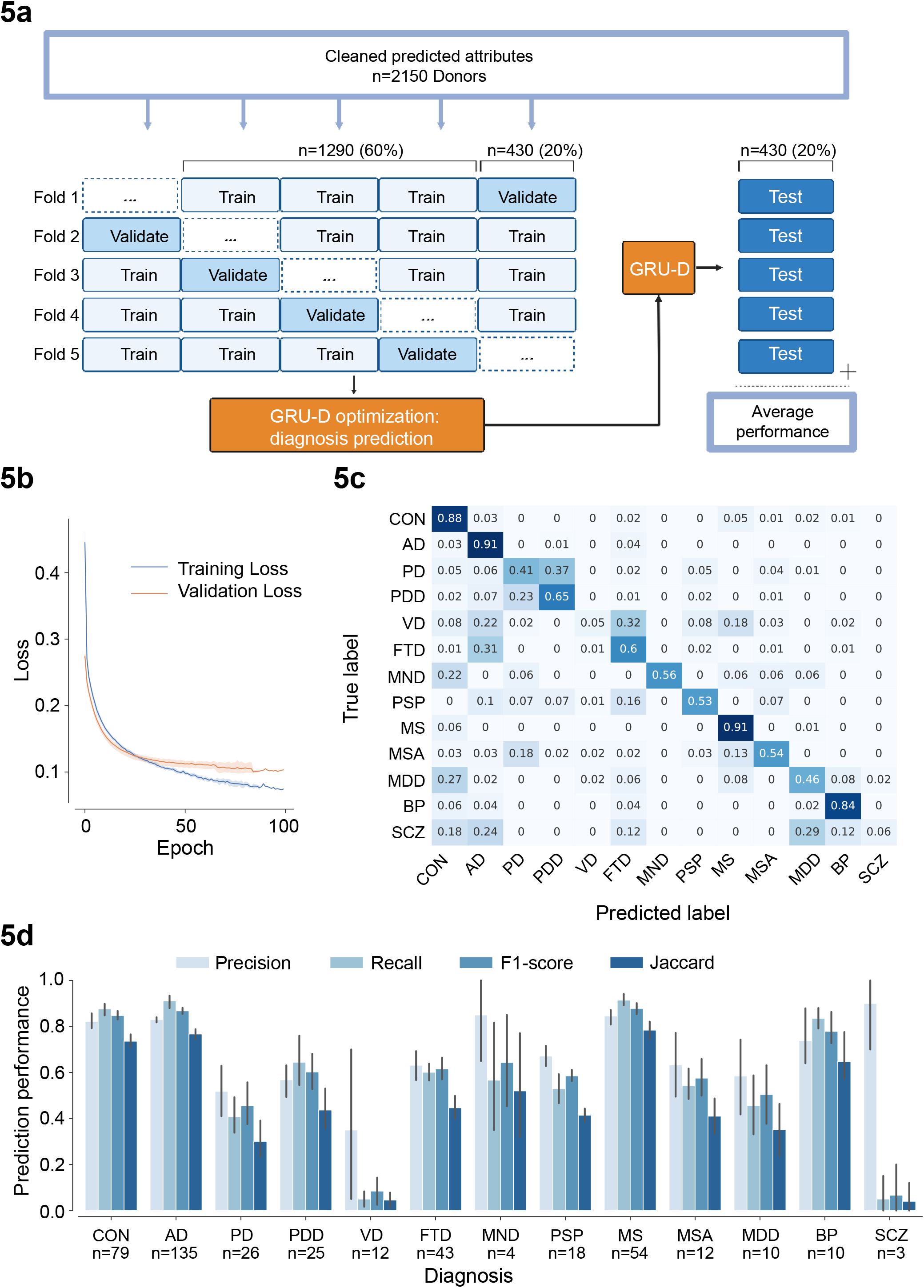
Predictive modeling of brain disorders from clinical signs and symptoms. A) Workflow of the predictive modeling approach. B) Training loss and validation loss of the GRU-D model plotted against the number of epochs, averaged over 5 folds. C) Confusion matrix showing the average proportion of brain disorders in the tests set predicted correctly by the GRU-D model. D) Average GRU-D prediction performance per disorder, as measured by precision, recall, F1-score and Jaccard index.

## Discussion

In this study, we aimed to identify temporal disease trajectories from brain donors that have received a wide range of Neuropathological Diagnoses. We identified 90 signs and symptoms (including 4 synonyms) within 14 groupings in 5 broad domains including a cognitive, psychiatric, sensory, motor, and general domain. We trained, optimized, and selected a high performing NLP model to identify these signs and symptoms in individual sentences. This resulted in a unique high-quality dataset that showed clear disease specific profiles. For example, patients with MS scored particularly high for ‘Muscular weakness’, and ‘Mobility problems’, which is in line with this severely debilitating condition of the brain and spinal cord. Donors who received a Neuropathological Diagnosis associated with dementia were also highly frequently characterized by signs of ‘Dementia’, ‘Memory impairment’, ‘Language impairment’ according to the NLP models, while patients with FTD also showed pronounced enrichments for ‘changed behavior and personality’. Many of these signs and symptoms showed observational, temporal and survival profiles that differed between major brain disorders, potentially allowing for differential diagnosis. Many of these temporal and survival profiles of signs and symptoms also differed between fine-grained neurological diagnoses such as rare and complex dementias, alpha-synucleinopathies, FTD subtypes, and mental illnesses.

A rapidly increasing number of studies focuses on the relationship between neuropathology, and clinical manifestation of brain disorders both within and between disorders such as AD-subtypes^31^, complex and comorbid dementias^3^, FTD-subtypes^28^, alpha-synucleinopathies^6^ and mental illnesses^32^. The relationship between neuropathology, clinical syndromes, and associated signs and symptoms is often complex, in which many signs and symptoms can occur in many disorders, but the frequency and the temporal profiles of these signs and symptoms generally tends to differ between neuropathological conditions. Hence, signs and symptoms cannot function as fully objective indicators of the underlying neuropathology. However, they do allow for a probabilistic diagnosis, and are hence indicative of the underlying neuropathology. Currently, medical doctors largely perform this probabilistic task mentally, frequently resulting in erroneous diagnosis. With the advent of trainable Machine Learning models, new avenues to perform this probabilistic diagnosis task and contribute to the differential diagnosis process by medical specialists have opened. However, a cross disorder dataset with highly reliably defined Neuropathological Diagnosis was missing, which we have retrospectively generated here. Corroborating the notion that this dataset can contribute towards improved diagnosis of brain disorders from symptoms, we successfully implemented a recurrent neural network to predict the underlying Neuropathological Diagnosis from the most common diagnosis.

AD, DLB, FTD, and VD are the most common causes of dementia and as such also most frequently studied. However, in our cohort, we found that a significant number of patients with dementia suffer from rare and complex forms of dementia, which are much less studied. Interestingly, we found that most donors who were diagnosed with these rare and complex forms of dementia tend to manifest a sign of ‘Dementia’ at significantly later ages, but that their survival is shorter than the survival of donors with AD or FTD. Alpha-synucleinopathies with dementia also showed significantly shorter survival than AD and or FTD after the first sign of ‘Dementia’ was observed. These observations suggest that the underlying neuropathological mechanisms that contribute towards the manifestation of ‘Dementia’, might differ from the pathogenic mechanisms that result in death. These findings were in line with previous publications concerning the survival of patients with Dementia, in which patients with DLB had significantly shorter survival than patients with AD^14^. Our study also extends upon this work, since we were able to determine the survival profiles of a much wider range of dementias. Moreover, as our unique temporal datasets contained many signs and symptoms, we were able to integrate different signs and symptoms. For example, the temporal profile of ‘Dementia’ is different from the temporal profile of ‘Bradykinesia’, which was observed at considerable earlier ages.

Alpha-synucleinopathies are brain disorders that are characterized by alpha-synuclein protein aggregation, and include PD, PDD, DLB, and MSA. Typically, PD, without dementia, is characterized by extrapyramidal signs, with modest neuropsychiatric and cognitive difficulties, while PDD is characterized by more severe cognitive problems. DLB, on the contrary, is most prominently characterized by ‘Dementia’, with fewer or negligible extrapyramidal signs. Signs of ‘Dementia’ in the absence of extrapyramidal signs is also a common feature of AD, hence, DLB and AD are frequently misdiagnosed^12^. MSA, on the contrary, is characterized by severe motor difficulties, including extrapyramidal signs, without overt cognitive and or neuropsychiatric problems. These differential signs and symptoms were clearly corroborated by this study. Importantly, there is considerable debate whether these alpha-synucleinopathies are merely different manifestations of the same pathophysiological processes, in which the brain regions that are most affected determine the clinical manifestation or that there are qualitative differences in the underlying pathophysiology^6^. We identified clear differential temporal and survival responses, between these four conditions after ‘Bradykinesia’ was first observed, which manifested at a considerably younger age in MSA, and was associated with shorter survival in MSA, and DLB, compared to PD and PDD. These differential temporal profiles are in line with the hypothesis that there are qualitative differences in the pathogenesis between MSA, DLB and PD/PDD, but hint at no qualitative differences between PD and PDD.

FTD is a group of highly heterogeneous conditions, in which degeneration of the frontotemporal cortex is a common hallmark^27,28, Staffaroni, 2022^. Different familial mutations and neuropathological subtypes were identified. Interestingly, these different familial mutations and or neuropathological subtypes are associated with the same clinical phenotypes. For example, studies have identified a behavioral syndrome, and a language syndrome, which are both common in different FTD subtypes, again illustrating the complex relationship between clinical signs and symptoms and underlying neuropathology. Our unique dataset can be used to identify differential (temporal) features that might be associated with these syndromes. For example, we found that PSP was uniquely characterized by lower observations of the sign of ‘Dementia’ compared to any other subtype. But temporally, ‘PSP’ is the latest affected disorder in the FTD subtypes. Moreover, this subtype is characterized by signs of ‘Bradykinesia’ and many other extrapyramidal signs. The unique dataset we present here, could be used as a resource to facilitate other research that links FTD subtypes to clinical syndromes.

Unlike the other brain disorders, mental illnesses, or psychiatric conditions, are disorders that are not characterized by prominent neuropathological changes, but are primarily associated with dysregulated emotions, cognitions, and behavior. Increasing lines of evidence suggest that mental illnesses are not discrete categories but more extreme values on a continuum^29,32^. However, the underlying axes are still largely unknown. We suggest that the temporal manifestation of signs and symptoms in these disorders might better represent the underlying pathophysiological processes, as these signs and symptoms are the direct output of the dysregulated neuronal activity. Future research should focus on studying the molecular profiles associated with specific psychiatric signs and symptoms, highlighting the relevance of this work for future studies in the context of both psychiatric and neurodegenerative disorders.

NLP models are increasingly implemented on medical health care records for information extraction, representation learning and phenotyping^33^. We have compared multiple approaches, including state of the art T5 and Google BERT transformer models, in which Google PubMedBERT showed the highest model performance. However, the differences between these models were quite modest, with all tested NLP models reaching high prediction performance. Several other studies have similarly developed outcome prediction models based on NLP of electronic health care records to identify signs and symptoms associated with brain disorders. For example, Chase (2017) identified 50 MS associated attributes that were able to predict the onset of MS using clinical notes before the actual clinical diagnosis in 30% of the cases, suggesting that machine learning models have tremendous clinical potential^34^. Jackson et al., 2017, identified 50 psychiatric symptoms, grouped into 5 categories, associated with severe mental illnesses, and found that cases with severe mental illnesses exhibited these symptoms more frequently than cases with non-severe mental illnesses^35^. Tran et al., 2017 trained a Convolutional Neural Network to predict mental illnesses directly from text data, and showed reasonable success rates (micro-F1 score of 0.63) for 11 common mental disorders^36^. Our work extends upon these studies, as we have taken a cross-brain disorder approach focusing on neuropathologically defined disorders. This work suggests that temporal cross-disorder signs and symptoms can function as reliable predictors of brain disease, potentially even extending to rare and complex diseases.

This work has several limitations related to missing values, retroactively scoring observations, and NLP models, potentially resulting in multiple levels in which (mis) interpretation could have emerged. For example, we were not able to distinguish between headaches and migraines fully reliably, even though the neurobiological mechanisms are generally considered to be quite different. Moreover, the summaries were made upon all health care records that were obtained by the trained staff from the NBB. Some medical doctors were not able to provide us with information, hence not all files are of equal quality and depth, which should have resulted in some missing values. Labeling errors could have been made, and might have influenced some of the results. Retroactive inspection of the signs and symptoms is based upon interpretation of the descriptions, in which it is not always possible to completely reliably determine whether a sign or symptoms was observed, or a mere hint of a sign or symptoms. Additionally, signs and symptoms were identified in several iterations; however, it is likely that some important signs and symptoms for specific brain diseases were missed. Additionally, the definitions of the signs and symptoms might not always be fully compliant with other existing frameworks, as they were tuned towards the Dutch health care system, and the NBB summaries and records. Moreover, the differential findings concerning the temporal and survival profiles between Neuropathological Diagnosis, might be unrelated to the neuropathological status *per se*, and might be confounded by additional variables such as euthanasia, drug treatments, and both medical and neuropathological comorbidities. Lastly, the Neuropathological Diagnoses were given over long periods of time by different neuropathologists, with the aim of tissue dispensation, not for clinical diagnostic purposes, and might deviate from clinical diagnosis.

### Future perspectives

The dataset with sentences and labels is associated as a supplemental table such that machine learning researchers who aim to build NLP models to score signs and symptoms can include this information into their modeling projects. This dataset could also be very valuable for future studies that aim to perform predictive modeling of the underlying neuropathology using signs and symptoms, and for researchers that are looking for clinical and temporal markers to distinguish subtypes of disorders. Lastly, this resource can be used for survival analysis of brain disorders after specific signs or symptoms, or combinations thereof, were observed, with the aim of improved survival predictions.

## Data Availability

All data produced in the present study are available upon reasonable request to the authors

---

**Supplemental Figure 1.**
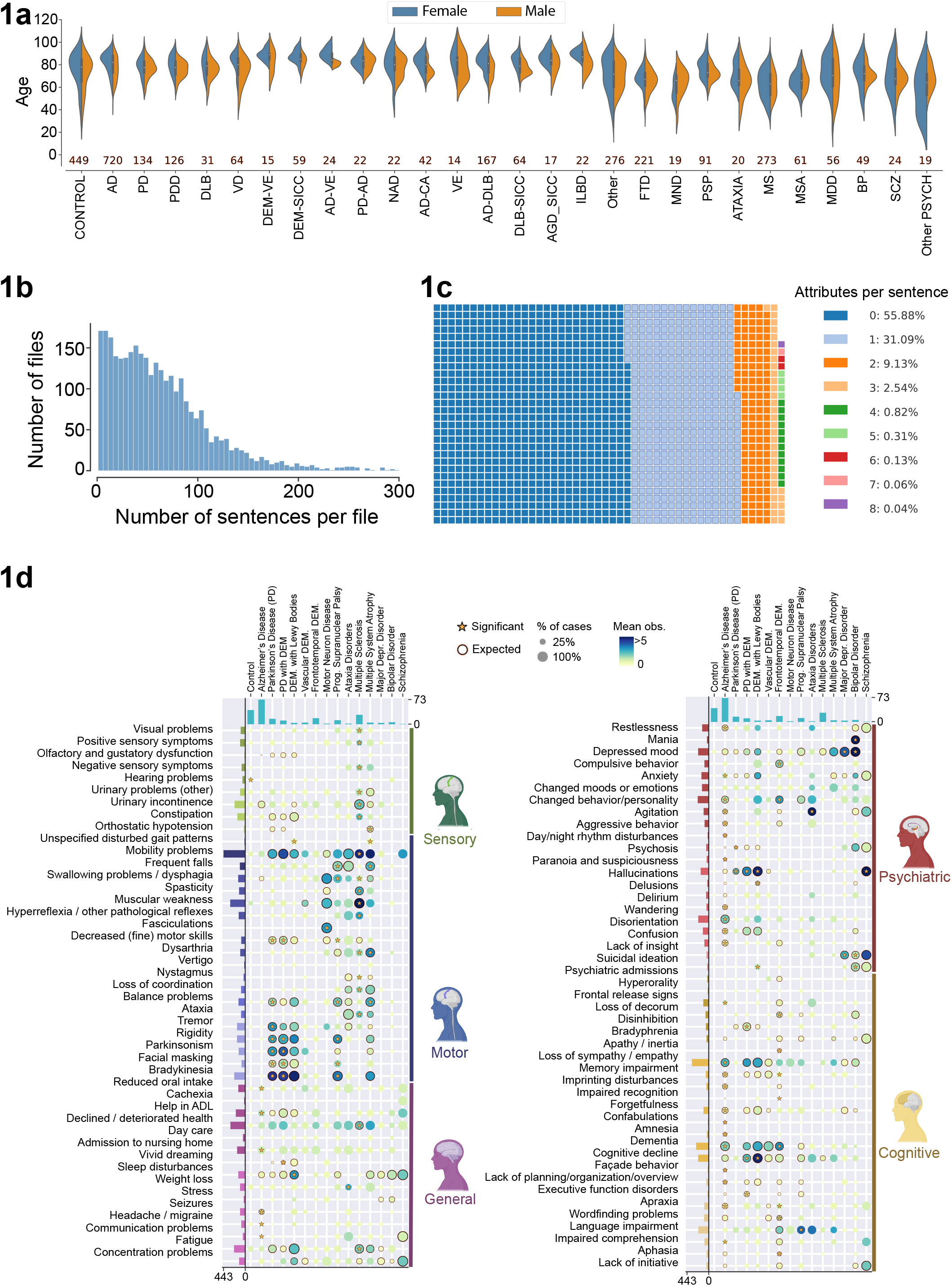
General outline of all the data. A) violin plot showing the age at death of the most common Neuropathological Diagnoses, separated by sex, with the number of donors in each category in the sample name. B) Histogram of the number of sentences in the ‘clinical history’ per donor file. C) Waffle chart of the number of labels per sentence for the labeled training dataset. D) Dot plot and significance testing of signs and symptoms across disorders for the labeled dataset. The dot plot contains information about the proportion of donors in which an attribute was observed, corresponding to the size of the dot. The mean number of observations corresponding to the color of the dot. Significance, asterisk, is based upon a permutation test, with an FDR corrected P < 0.1. Orange surrounding circles highlight signs and symptoms that, *a priori*, were medically expected to be observed given a diagnosis.

**Supplemental Figure 2.**
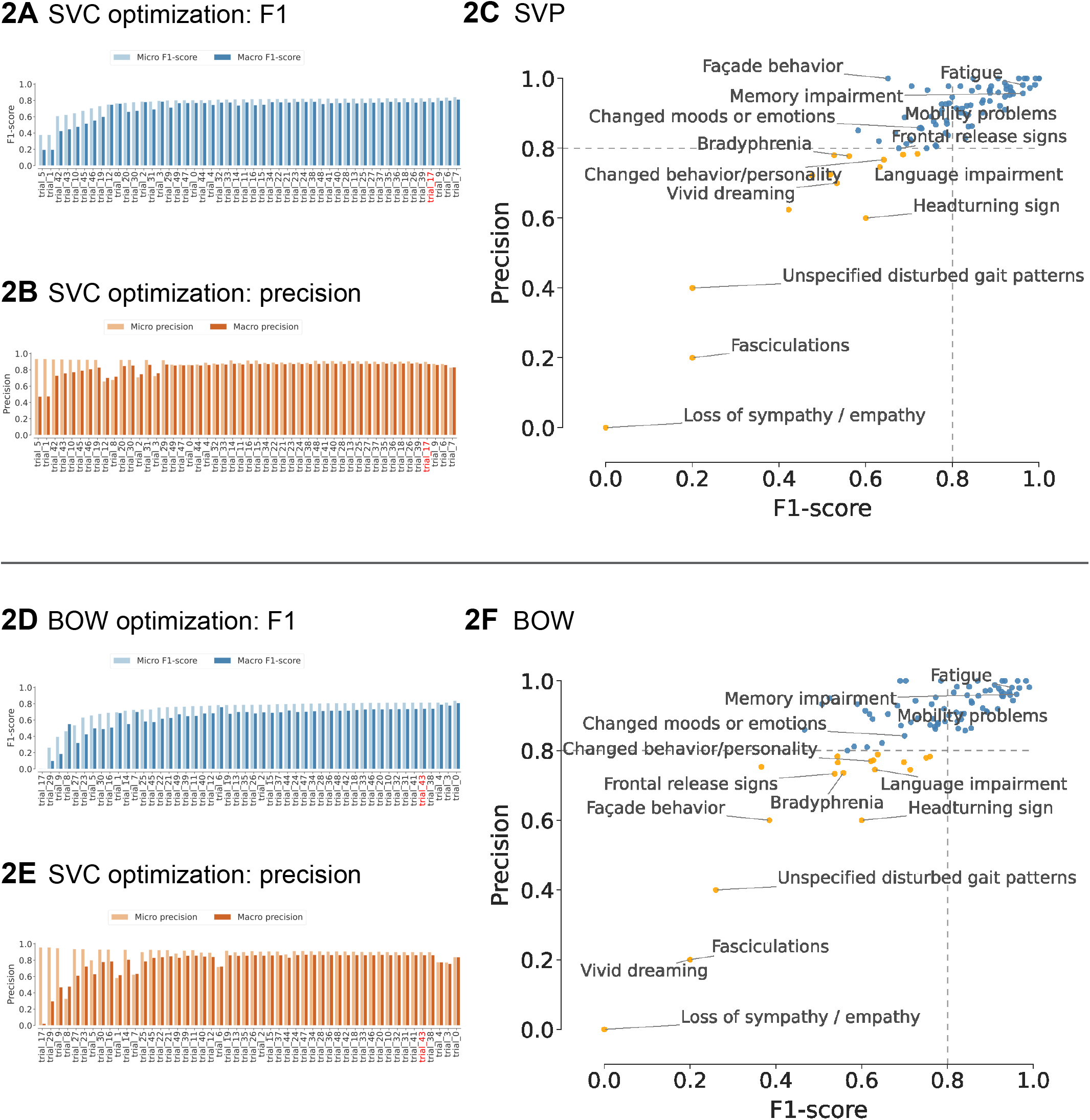
NLP model optimization for SVC, and BOW. A, B, D, E) Bar plot of micro (light blue) and macro (dark blue) F1 scores (A, D) and of micro (light orange) and macro (dark orange) precision (B, E) for different optimization trials from Optuna for SVC (A & B), BOW (D & E). C & F) scatterplot depicting the precision and F1-score for SVC (C) and BOW (F).

**Supplemental Figure 3.**
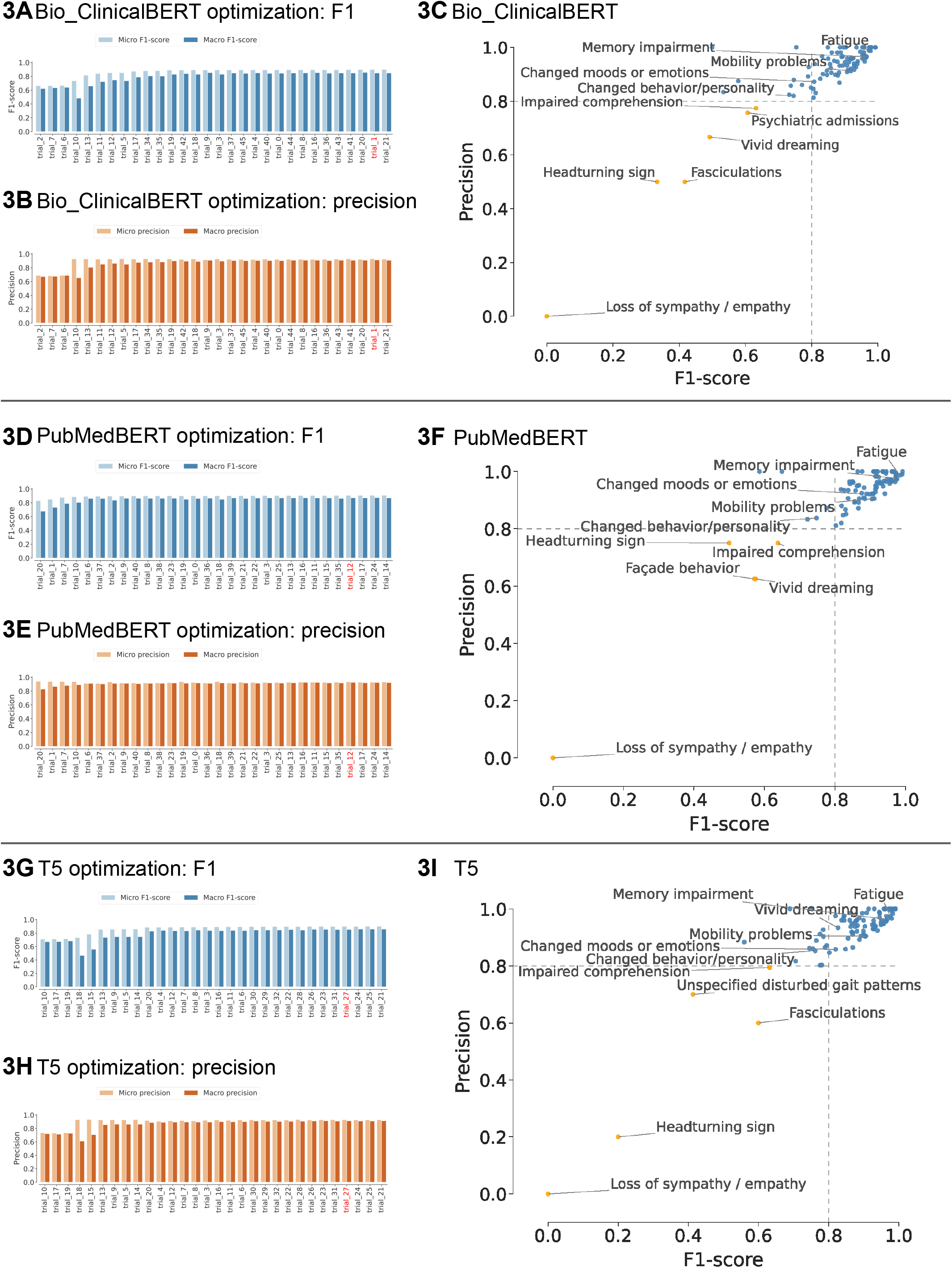
NLP model optimization for PubMedBERT, Bio_ClinicalBERT, and T5. A, B, D, E, G, H) Bar plot of micro (light blue) and macro (dark blue) F1 scores (A, D, G) and of micro (light orange) and macro (dark orange) precision (B, E, H) for different optimization trials from Optuna for PubMedBERT (A & B), Bio_ClinicalBERT (D & E) and T5 (G, H). C, F, I) scatterplot depicting the precision and F1-score for PubMedBERT (C), Bio_ClinicalBERT (F), and T5 (F).

**Supplemental Figure 4.**
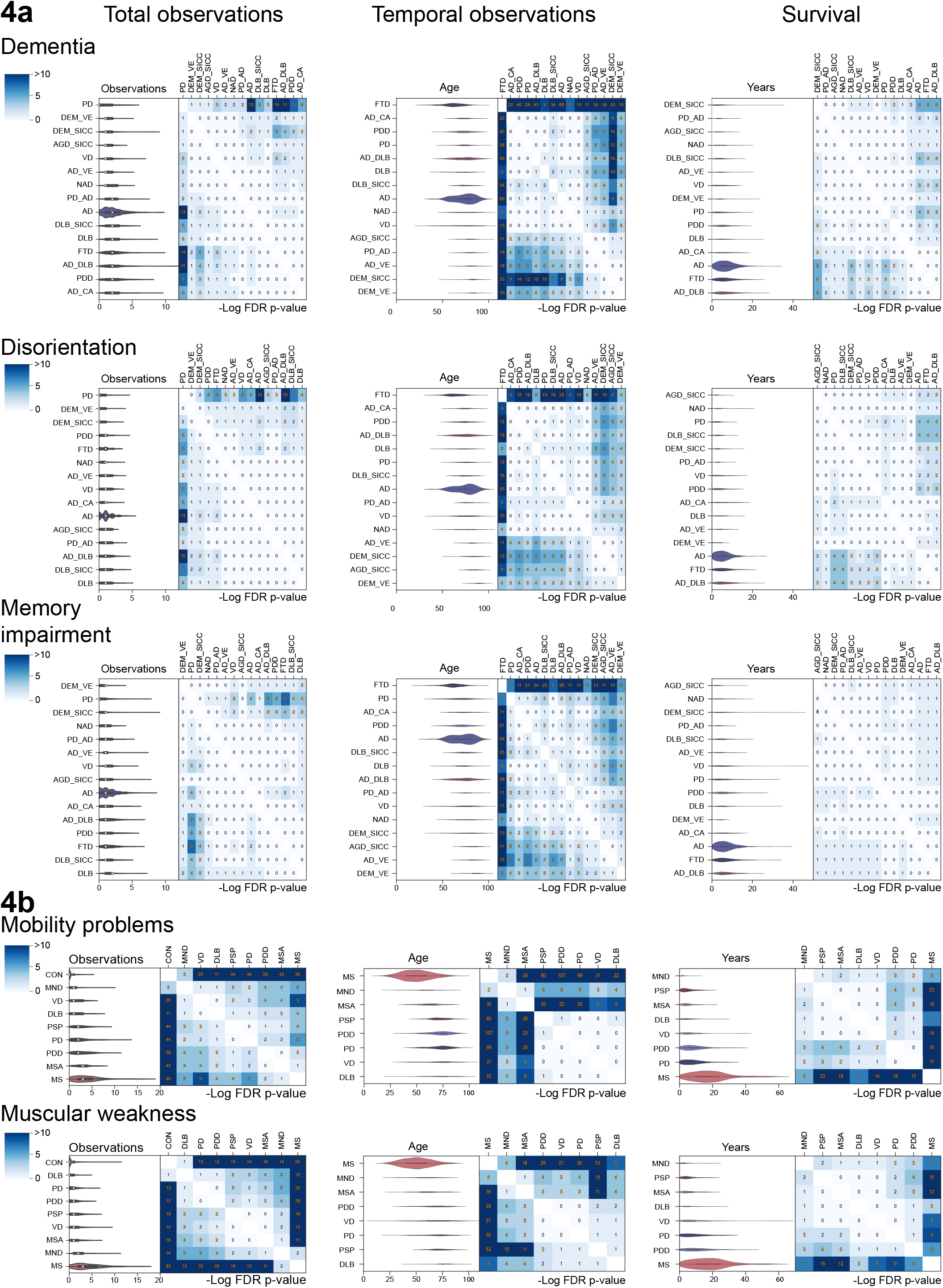
Observation, temporal and survival distributions across signs and symptoms. A) Violin plots depicting the observation distributions, the temporal distribution, and survival distributions of Dementias, using ‘Dementia’, ‘Disorientation’ and ‘Memory Impairment’ for dementias with different causes, including rare and complex dementias. Each violin plot is accompanied by a heatmap showing the results of pairwise significance testing, with -10log FDR corrected p-values depicted in orange when significant (p <= 0.01). B) Violin plots with heatmap depicting the observation distributions, the temporal distribution, and survival distributions of motor symptoms ‘Mobility Problems’, ‘Muscular Weakness’.

**Supplemental Figure 5.**
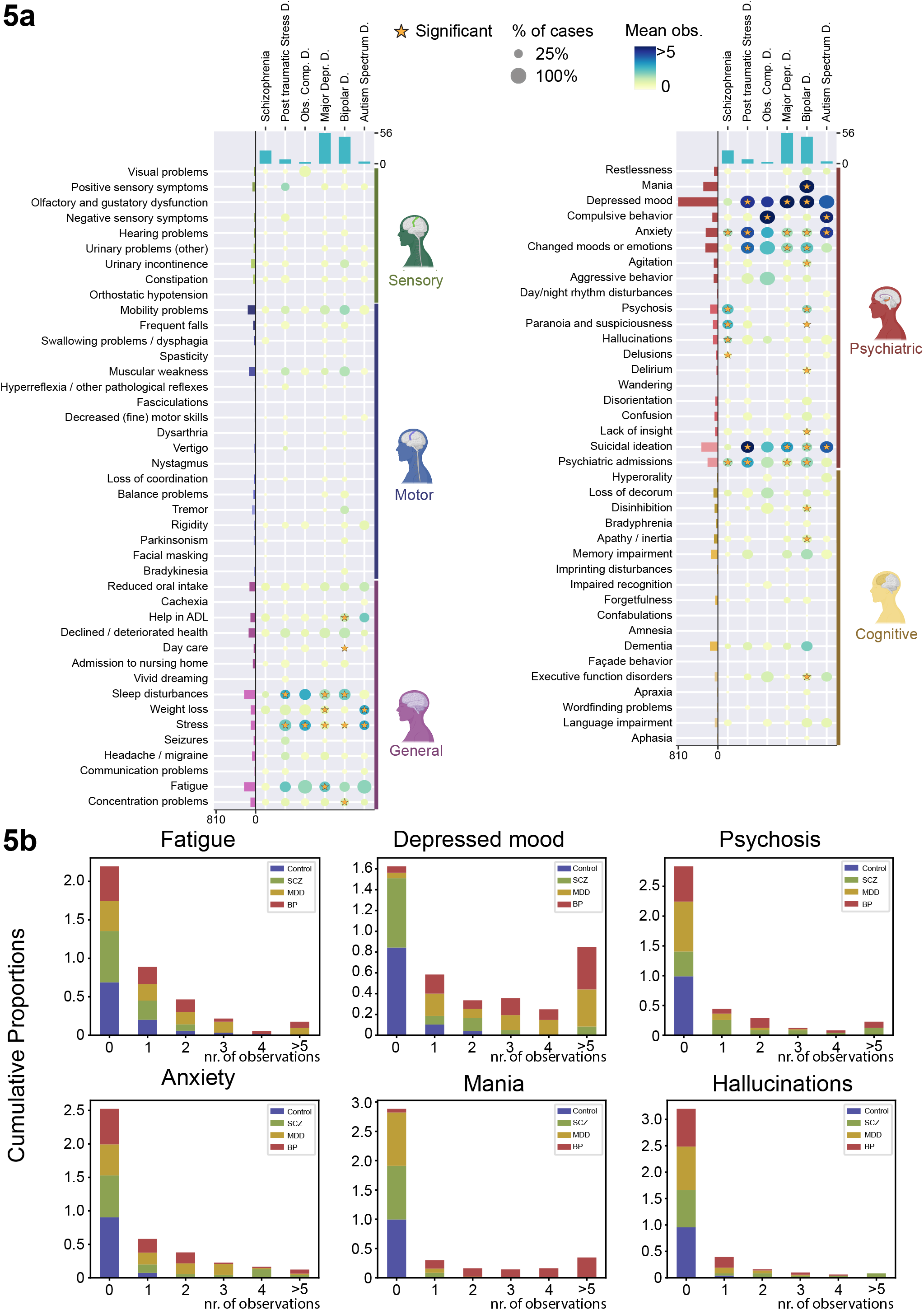
Analysis of mental illnesses. A) Dot plot and significance testing of signs and symptoms across mental illnesses. B) Stacked bar plot showing cumulative proportions of 3 major mental illnesses (SCZ, BP, MDD and controls) across the number of year observations from 0 to 5, in which donors with more than 5-year observations were merged into the 5 or more years category. Stacked bar plots were made for ‘Fatigue’, ‘Depressed mood’, ‘Psychosis’, ‘Anxiety’, ‘Mania’, ‘Hallucinations’.

## Acknowledgements

We would like to express our gratitude to the ‘Stichting Vrienden van het Herseninstituut’ for their financial support that made this work possible. We thank Dr. Marta Scarioni for checking and validating the neurological validity of the identified signs and symptoms. We also want to show our gratitude towards all the individuals who have decided to become a donor for the NBB and their family members for this act of courage.

